# Application of severity classification after return of spontaneous circulation for predicting long-term outcomes in cardiac arrest survivors

**DOI:** 10.1101/2025.11.03.25339449

**Authors:** Jr-Jiun Lin, Chien-Hua Huang, Wei-Ting Chen, Hsin-Yu Lee, Wen-Jone Chen, Wei-Tien Chang, Hooi-Nee Ong, Min-Shan Tsai

## Abstract

**Background:** Accurately predicting outcomes for cardiac arrest (CA) survivors is critical, and several severity scores have been developed to predict outcomes at hospital discharge. However, the ability of these scores to predict long-term outcomes at 6 months and 1 year in CA survivors remains unexplored.

**Methods:** This retrospective observational study enrolled 1773 adult patients with nontraumatic CA in the emergency department of National Taiwan University Hospital between January 2011 and June 2023 without interhospital transfer. Data on clinical variables, resuscitation events, post-arrest care, and outcomes were collected, and the disease severity was classified using the sCAHP, rCAST, and TIMECARD scores. The outcomes included 6-month and 1-year survival. The predictive ability of these scores was evaluated using receiver operating characteristic curves.

**Results:** Among the 1773 enrolled patients, 501 (28.3%) survived to discharge, 361 (20.4%) survived for 6 months, and 346 (19.5%) survived for 1 year. The area under the receiver operating characteristic curves (AUCs) for 6-month survival were 0.787, 0.733, and 0.810 for the sCAHP, rCAST, and TIMECARD scores, respectively. For 1-year survival, the AUC values were 0.788, 0.734, and 0.816, respectively. The sCAHP and TIMECARD scores demonstrated better discriminatory performance than did the rCAST score for both 6-month and 1-year survival. Compared with the sCAHP and rCAST scores, the TIMECARD score exhibited superior discriminatory ability for 1-year survival, particularly in younger patients (age ≤65 years), those with in-hospital CA (IHCA), and those with good pre-arrest neurological status.

**Conclusion:** The sCAHP, rCAST, and TIMECARD scores demonstrated good discrimination and calibration in predicting long-term outcomes in CA survivors. The TIMECARD score exhibited better discriminatory performance for 1-year survival, particularly in the subgroup of younger age, IHCA, and good pre-arrest neurological function.

## Introduction

The economic and health-care burden associated with acute critical care and long-term management of cardiac arrest (CA) survivors after the return of spontaneous circulation (ROSC) is substantial for both patients’ families and the community. Evaluating illness severity after CA is crucial for guiding post-arrest care strategies, including targeted temperature management (TTM) (1–3). Thus, accurately classifying illness severity and predicting clinical outcomes in successfully resuscitated patients are essential.

Various severity scores have been developed, including the Cardiac Arrest Hospital Prognosis (CAHP) score (4), the post-Cardiac Arrest Syndrome for induced Therapeutic hypothermia (CAST) score (5), and the TaIwan network of targeted temperature ManagEment for CARDiac arrest (TIMECARD) score (6) published by our study group. Some parameters in these severity scores are difficult to obtain. Thus, the simplified CAHP (sCAHP) score (7–9), which omits the no-flow interval, and the revised CAST (rCAST) score (9, 10), which excludes the gray matter to white matter attenuation ratio, serum albumin, and hemoglobin, have been further refined for improved clinical applicability.

With advancements in interventions, monitoring, and treatments during the post-cardiac arrest period, patients’ survival and neurological outcomes have correspondingly improved over the past decades (11, 12). Thus, predicting long-term outcomes has become increasingly crucial for CA survivors. However, most severity scores have been developed to predict short-term outcomes, whereas long-term prognosis, such as survival at 6 months, 1 year, or beyond, remains less studied (13–15). Few studies have evaluated long-term outcomes in CA survivors by using established severity scores (16, 17), but these have been limited by small sample sizes or inadequate follow-up periods. Moreover, most studies on long-term outcomes have focused on patients with out-of-hospital cardiac arrest (OHCA), whereas studies on in-hospital cardiac arrest (IHCA) or Asian populations remain limited. A previous study (18) recommended that 7 days of observation in neurologic recovery for comatose cardiac arrest survivors, with minimal likelihood of further long-term improvement. However, whether these individuals have a higher post-discharge mortality rate over the long term remains a question of interest for their families. Therefore, the present study evaluated the ability of published severity scores, including the sCAHP, rCAST, and TIMECARD scores, to predict long-term survival in CA survivors in Taiwan.

## Materials and Methods

### Study design and patient enrollment

This retrospective single-center cohort study was conducted in the emergency department (ED) of National Taiwan University Hospital (NTUH), a 2500-bed medical center in Taipei City (19), which has a population of 2.68 million and covers an area of 272 square kilometers. This study was approved by the Institutional Review Board (IRB) of NTUH (IRB number: 202407049RIND), and the requirement for informed consent was waived.

There were 2461 consecutive adult patients with non-traumatic CA who successfully achieved ROSC in the ED of NTUH between January 2011 and June 2023 were initially identified. Patients who were traumatic, pregnant, younger than 18 years of age, transferred to or from another hospital, registered in TIMECARD registry, or lost to follow-up within 1 year were excluded. Thus, a total of 1773 adult patients with non-traumatic CA were finally included (Figure 1).

**Figure 1.**
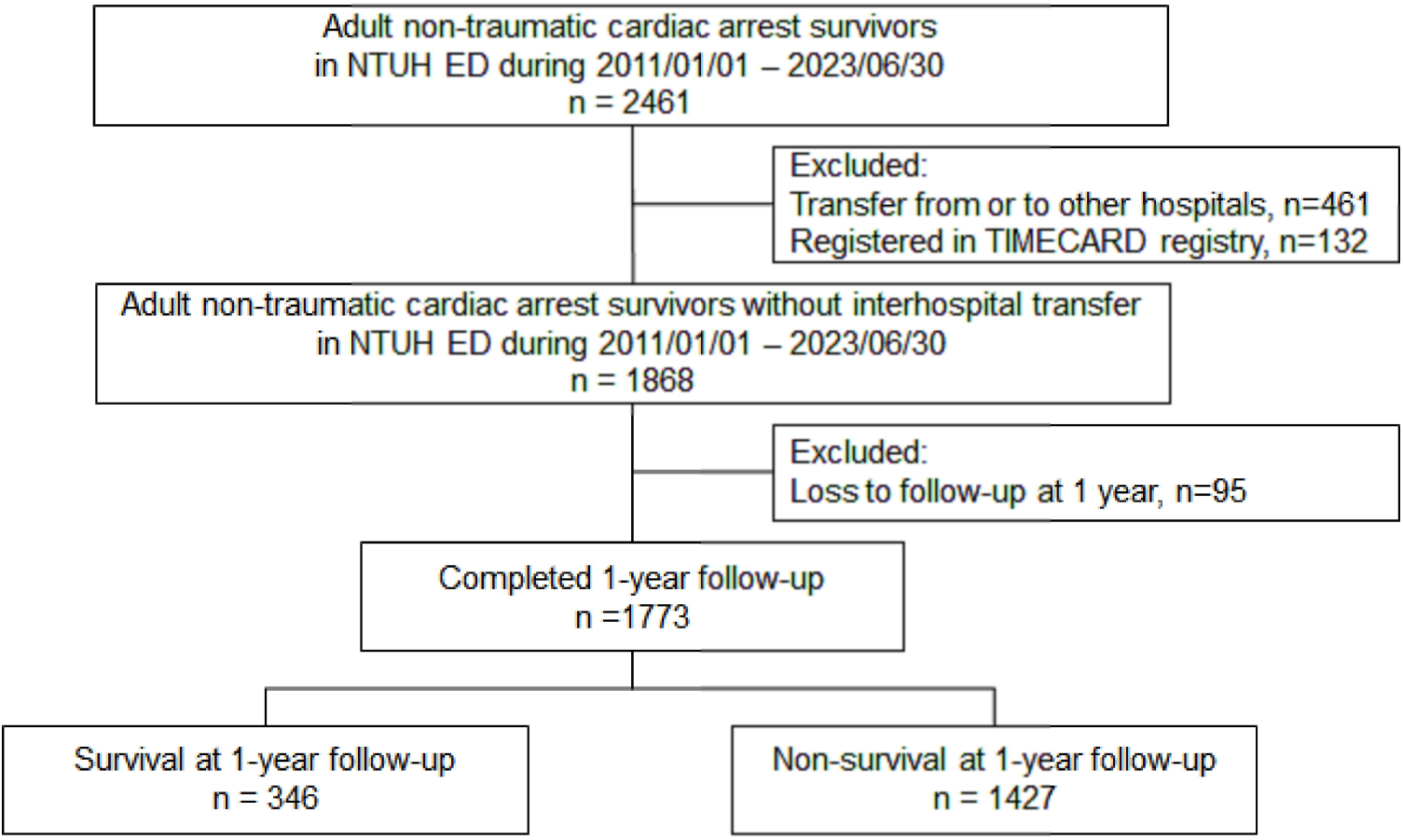
The flow chart of patient enrollment. ED: emergency department; NTUH: National Taiwan University Hospital; TIMECARD registry: TaIwan network of targeted temperature ManagEment for CARDiac arrest (TIMECARD) registry.

### Data collection

Patient data regarding baseline characteristics, underlying medical conditions, resuscitation events, and post-arrest care were collected from medical and emergency medical services records using predesigned questionnaires. Malignancy was defined as stable or active cancer (6). Pre-arrest neurological status was evaluated by primary care physicians who were blinded to the current study, using Glasgow–Pittsburgh cerebral performance category (CPC) scores based on previous medical records or statements from the patient’s family. A CPC score of 1 (good performance) or 2 (moderate disability) indicated good neurological status, whereas a CPC score of 3 (severe disability), 4 (vegetative state), or 5 (death) indicated poor neurological status (20). Prehospital ROSC was defined as the return of a heartbeat with palpable pulsation and obtainable peripheral blood pressure before hospital arrival in OHCA survivors (11, 12). A shockable rhythm was defined as pulseless ventricular tachycardia or ventricular fibrillation during resuscitation, and defibrillation was delivered via automated external defibrillators or manual defibrillators (11, 12). The call-to-start cardiopulmonary resuscitation (CPR) duration was the interval between the call for help and the start of chest compressions (6, 21). The CPR duration was defined as the time from the initiation of chest compressions to ROSC, including both prehospital and in-hospital periods (6, 21). The blood gas analyses, pH values, and lactate levels were obtained from the first blood sample collected after CA. TTM was performed when clinically indicated in comatose CA survivors (21). Extracorporeal CPR was recorded when performed as indicated during the index resuscitation. Emergent coronary angiography was documented when performed within 24 hours after ROSC (22). Severity scores used to predict outcomes—the sCAHP (4, 7–9), rCAST (5, 9, 10), and TIMECARD score (6)—were calculated for all enrolled patients (Supplementary Table 1).

### Outcome measurements

The primary outcome of this study was 1-year survival, and the secondary outcomes were survival-to-discharge, neurological recovery at discharge, and 6-month survival. Long-term follow-up assessments were conducted by clinical staff of the discharging ward using structured telephone questionnaires, particularly at 6 months and 1 year following ROSC. Neurological outcomes at discharge were evaluated by duty physicians and classified as favorable for patients with a CPC score of 1 or 2 and unfavorable for those with a CPC score of 3, 4, or 5.

### Statistical analysis

Continuous variables were presented as mean ± standard deviation, and the differences between groups were analyzed using the independent t-test. Categorical variables were presented as numbers and percentages, and the differences between groups were assessed using the chi-squared test. The discriminatory ability of the severity scores was evaluated using the area under the receiver operating characteristic curve (AUC), and pairwise AUC comparisons were performed utilizing the method established by DeLong et al (23). The calibration of the severity scores was analyzed using the Hosmer-Lemeshow goodness-of-fit test (24). Statistical significance was set at *p* < 0.05. All statistical analyses were performed using Statistical Package for Social Sciences Statistics 21.0 (IBM Corp., Armonk, N.Y., USA), except the DeLong tests, which were performed using MedCalc Statistical Software version 18.2.1 (MedCalc Software bvba, Ostend, Belgium).

## Results

Among the 1773 patients who were successfully resuscitated, 501 (28.3%) survived to discharge, with 384 (21.7%) exhibiting favorable neurological outcomes at discharge, 361 (20.4%) surviving at the 6-month follow-up, and 346 patients (19.5%) survived at the 1-year follow-up (Figure 1). The average age of enrolled patients was 67.81 ± 15.78 years, and 1147 (64.7%) patients were men. The baseline characteristics, resuscitation events, post-arrest care, outcomes, and severity scores between the groups stratified by 1-year survival are presented in Table 1.

**Table 1.**
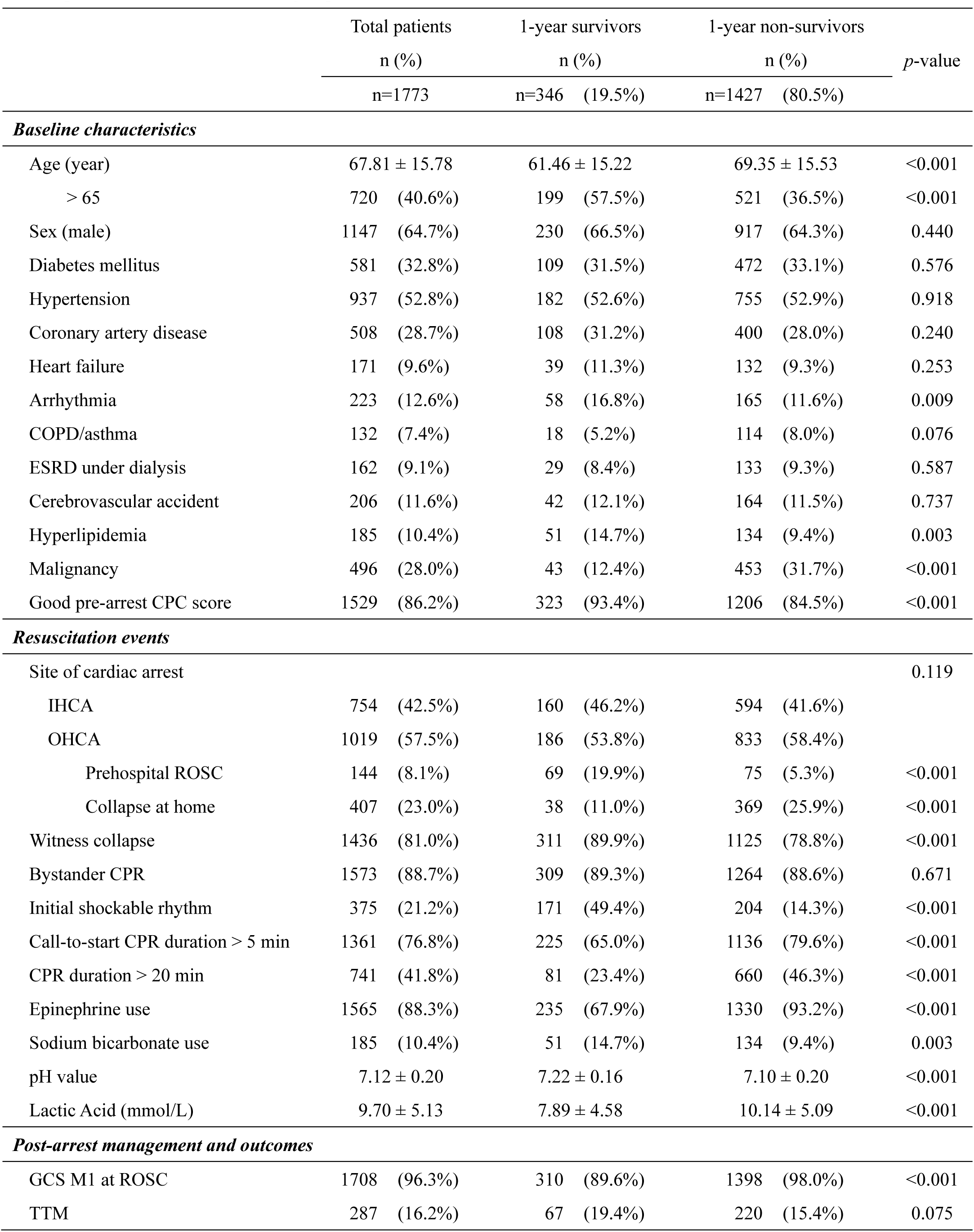
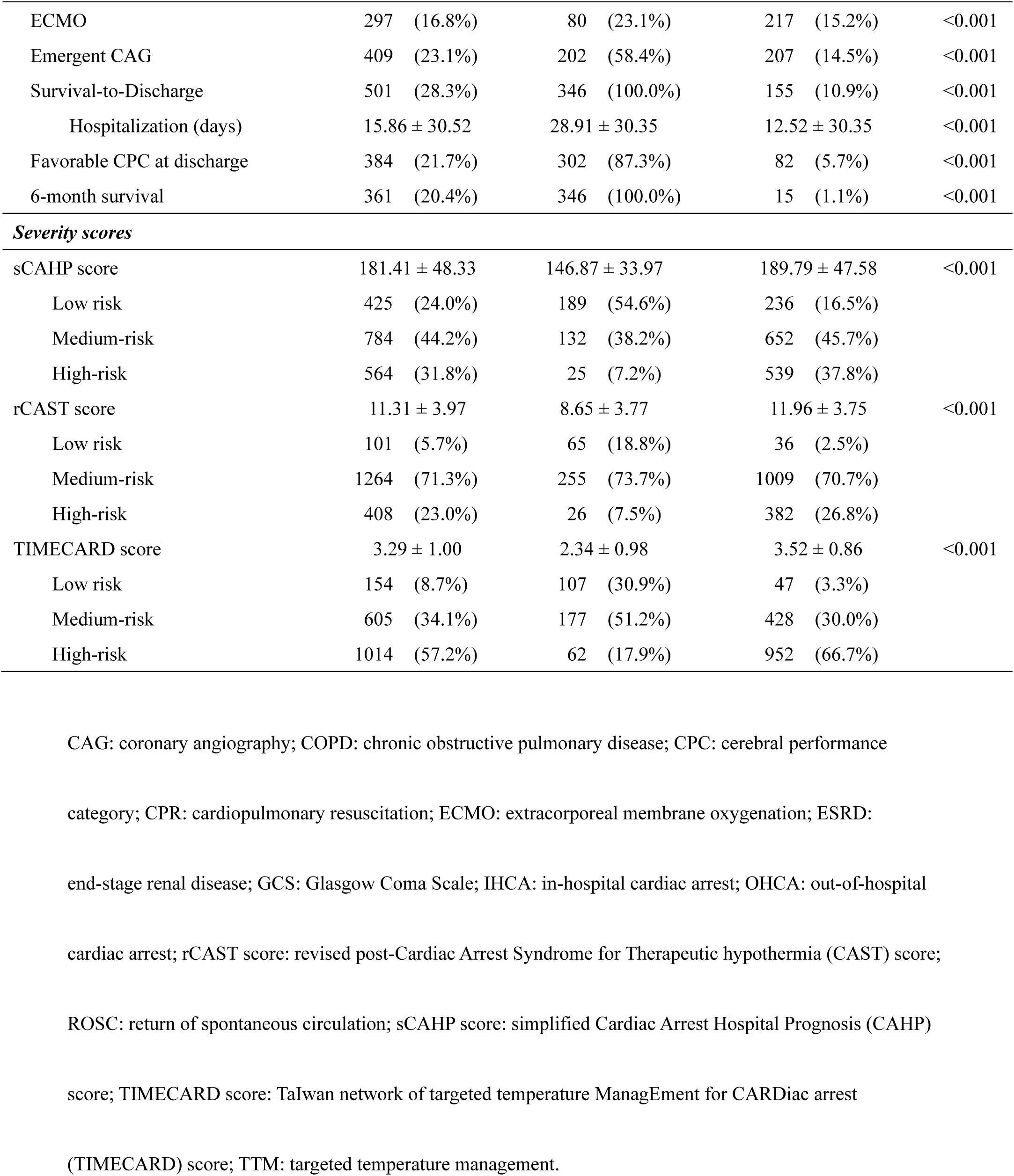
Baseline characteristics, resuscitation events, post-arrest management, outcomes, and prediction scores between the groups.

|  | Total patients |  | 1-year survivors |  | 1-year non-survivors |  | p-value |
| --- | --- | --- | --- | --- | --- | --- | --- |
|  | n (%) |  | n (%) |  | n (%) |  |  |
|  | n=1773 |  | n=346 (19.5%) |  | n=1427 (80.5%) |  |  |
| **Baseline characteristics** |  |  |  |  |  |  |  |
| Age (year) | 67.81 ± 15.78 |  | 61.46 ± 15.22 |  | 69.35 ± 15.53 |  | <0.001 |
| > 65 | 720 | (40.6%) | 199 | (57.5%) | 521 | (36.5%) | <0.001 |
| Sex (male) | 1147 | (64.7%) | 230 | (66.5%) | 917 | (64.3%) | 0.440 |
| Diabetes mellitus | 581 | (32.8%) | 109 | (31.5%) | 472 | (33.1%) | 0.576 |
| Hypertension | 937 | (52.8%) | 182 | (52.6%) | 755 | (52.9%) | 0.918 |
| Coronary artery disease | 508 | (28.7%) | 108 | (31.2%) | 400 | (28.0%) | 0.240 |
| Heart failure | 171 | (9.6%) | 39 | (11.3%) | 132 | (9.3%) | 0.253 |
| Arrhythmia | 223 | (12.6%) | 58 | (16.8%) | 165 | (11.6%) | 0.009 |
| COPD/asthma | 132 | (7.4%) | 18 | (5.2%) | 114 | (8.0%) | 0.076 |
| ESRD under dialysis | 162 | (9.1%) | 29 | (8.4%) | 133 | (9.3%) | 0.587 |
| Cerebrovascular accident | 206 | (11.6%) | 42 | (12.1%) | 164 | (11.5%) | 0.737 |
| Hyperlipidemia | 185 | (10.4%) | 51 | (14.7%) | 134 | (9.4%) | 0.003 |
| Malignancy | 496 | (28.0%) | 43 | (12.4%) | 453 | (31.7%) | <0.001 |
| Good pre-arrest CPC score | 1529 | (86.2%) | 323 | (93.4%) | 1206 | (84.5%) | <0.001 |
| **Resuscitation events** |  |  |  |  |  |  |  |
| Site of cardiac arrest |  |  |  |  |  |  | 0.119 |
| IHCA | 754 | (42.5%) | 160 | (46.2%) | 594 | (41.6%) |  |
| OHCA | 1019 | (57.5%) | 186 | (53.8%) | 833 | (58.4%) |  |
| Prehospital ROSC | 144 | (8.1%) | 69 | (19.9%) | 75 | (5.3%) | <0.001 |
| Collapse at home | 407 | (23.0%) | 38 | (11.0%) | 369 | (25.9%) | <0.001 |
| Witness collapse | 1436 | (81.0%) | 311 | (89.9%) | 1125 | (78.8%) | <0.001 |
| Bystander CPR | 1573 | (88.7%) | 309 | (89.3%) | 1264 | (88.6%) | 0.671 |
| Initial shockable rhythm | 375 | (21.2%) | 171 | (49.4%) | 204 | (14.3%) | <0.001 |
| Call-to-start CPR duration > 5 min | 1361 | (76.8%) | 225 | (65.0%) | 1136 | (79.6%) | <0.001 |
| CPR duration > 20 min | 741 | (41.8%) | 81 | (23.4%) | 660 | (46.3%) | <0.001 |
| Epinephrine use | 1565 | (88.3%) | 235 | (67.9%) | 1330 | (93.2%) | <0.001 |
| Sodium bicarbonate use | 185 | (10.4%) | 51 | (14.7%) | 134 | (9.4%) | 0.003 |
| pH value | 7.12 ± 0.20 |  | 7.22 ± 0.16 |  | 7.10 ± 0.20 |  | <0.001 |
| Lactic Acid (mmol/L) | 9.70 ± 5.13 |  | 7.89 ± 4.58 |  | 10.14 ± 5.09 |  | <0.001 |
| **Post-arrest management and outcomes** |  |  |  |  |  |  |  |
| GCS M1 at ROSC | 1708 | (96.3%) | 310 | (89.6%) | 1398 | (98.0%) | <0.001 |
| TTM | 287 | (16.2%) | 67 | (19.4%) | 220 | (15.4%) | 0.075 |

|  |  |  |  |  |
| --- | --- | --- | --- | --- |
| ECMO | 297 (16.8%) | 80 (23.1%) | 217 (15.2%) | <0.001 |
| Emergent CAG | 409 (23.1%) | 202 (58.4%) | 207 (14.5%) | <0.001 |
| Survival-to-Discharge | 501 (28.3%) | 346 (100.0%) | 155 (10.9%) | <0.001 |
| Hospitalization (days) | 15.86 ± 30.52 | 28.91 ± 30.35 | 12.52 ± 30.35 | <0.001 |
| Favorable CPC at discharge | 384 (21.7%) | 302 (87.3%) | 82 (5.7%) | <0.001 |
| 6-month survival | 361 (20.4%) | 346 (100.0%) | 15 (1.1%) | <0.001 |
| **Severity scores** |  |  |  |  |
| sCAHP score | 181.41 ± 48.33 | 146.87 ± 33.97 | 189.79 ± 47.58 | <0.001 |
| Low risk | 425 (24.0%) | 189 (54.6%) | 236 (16.5%) |  |
| Medium-risk | 784 (44.2%) | 132 (38.2%) | 652 (45.7%) |  |
| High-risk | 564 (31.8%) | 25 (7.2%) | 539 (37.8%) |  |
| rCAST score | 11.31 ± 3.97 | 8.65 ± 3.77 | 11.96 ± 3.75 | <0.001 |
| Low risk | 101 (5.7%) | 65 (18.8%) | 36 (2.5%) |  |
| Medium-risk | 1264 (71.3%) | 255 (73.7%) | 1009 (70.7%) |  |
| High-risk | 408 (23.0%) | 26 (7.5%) | 382 (26.8%) |  |
| TIMECARD score | 3.29 ± 1.00 | 2.34 ± 0.98 | 3.52 ± 0.86 | <0.001 |
| Low risk | 154 (8.7%) | 107 (30.9%) | 47 (3.3%) |  |
| Medium-risk | 605 (34.1%) | 177 (51.2%) | 428 (30.0%) |  |
| High-risk | 1014 (57.2%) | 62 (17.9%) | 952 (66.7%) |  |

CAG: coronary angiography; COPD: chronic obstructive pulmonary disease; CPC: cerebral performance
category; CPR: cardiopulmonary resuscitation; ECMO: extracorporeal membrane oxygenation; ESRD:
end-stage renal disease; GCS: Glasgow Coma Scale; IHCA: in-hospital cardiac arrest; OHCA: out-of-hospital
cardiac arrest; rCAST score: revised post-Cardiac Arrest Syndrome for Therapeutic hypothermia (CAST) score;
ROSC: return of spontaneous circulation; sCAHP score: simplified Cardiac Arrest Hospital Prognosis (CAHP)
score; TIMECARD score: TaIwan network of targeted temperature ManagEment for CARDIac arrest
(TIMECARD) score; TTM: targeted temperature management.

Compared with 1-year non-survivors, 1-year survivors had a younger age, higher incidences of arrhythmia and hyperlipidemia, a lower prevalence of malignancy, and a better pre-arrest neurological status. Patients with long-term survival had a lower incidence of CA occurring at home, higher incidences of witnessed collapses, more frequent initial shockable rhythms, shorter CPR durations, decreased epinephrine or sodium bicarbonate use, pH levels closer to neutral, lower lactic acid levels, and a lower prevalence of a Glasgow coma scale motor score of 1 at ROSC.

The receiver operating characteristic (ROC) curves for enrolled patients are illustrated in Figure 2. The AUC values of the sCAHP, rCAST, and TIMECARD scores were 0.767, 0.713, and 0.790 for survival-to-discharge; 0.804, 0.736, and 0.814 for favorable neurological outcomes at discharge; 0.787, 0.733, and 0.810 for 6-month survival; and 0.788, 0.734, and 0.816 for 1-year survival, respectively. The results of pairwise AUC comparisons performed using the DeLong tests are presented in Figure 2. The TIMECARD score demonstrated superior discriminatory performance for 6-month survival compared with the rCAST score (difference between AUC = 0.077, 95% confidence interval [CI] = 0.046 – 0.108, *p* < 0.001), and for 1-year survival compared with both the sCAHP (difference between AUC = 0.028, 95% CI = -0.001 – 0.057, *p* = 0.055) and rCAST scores (difference between AUC = 0.082, 95% CI = 0.051 – 0.113, *p* < 0.001). The calibration of the severity scores in predicting long-term outcomes was evaluated using the Hosmer-Lemeshow goodness-of-fit test (Supplementary Figure 1), and the results revealed no statistically significant differences between the predicted and observed outcomes.

**Figure 2.**
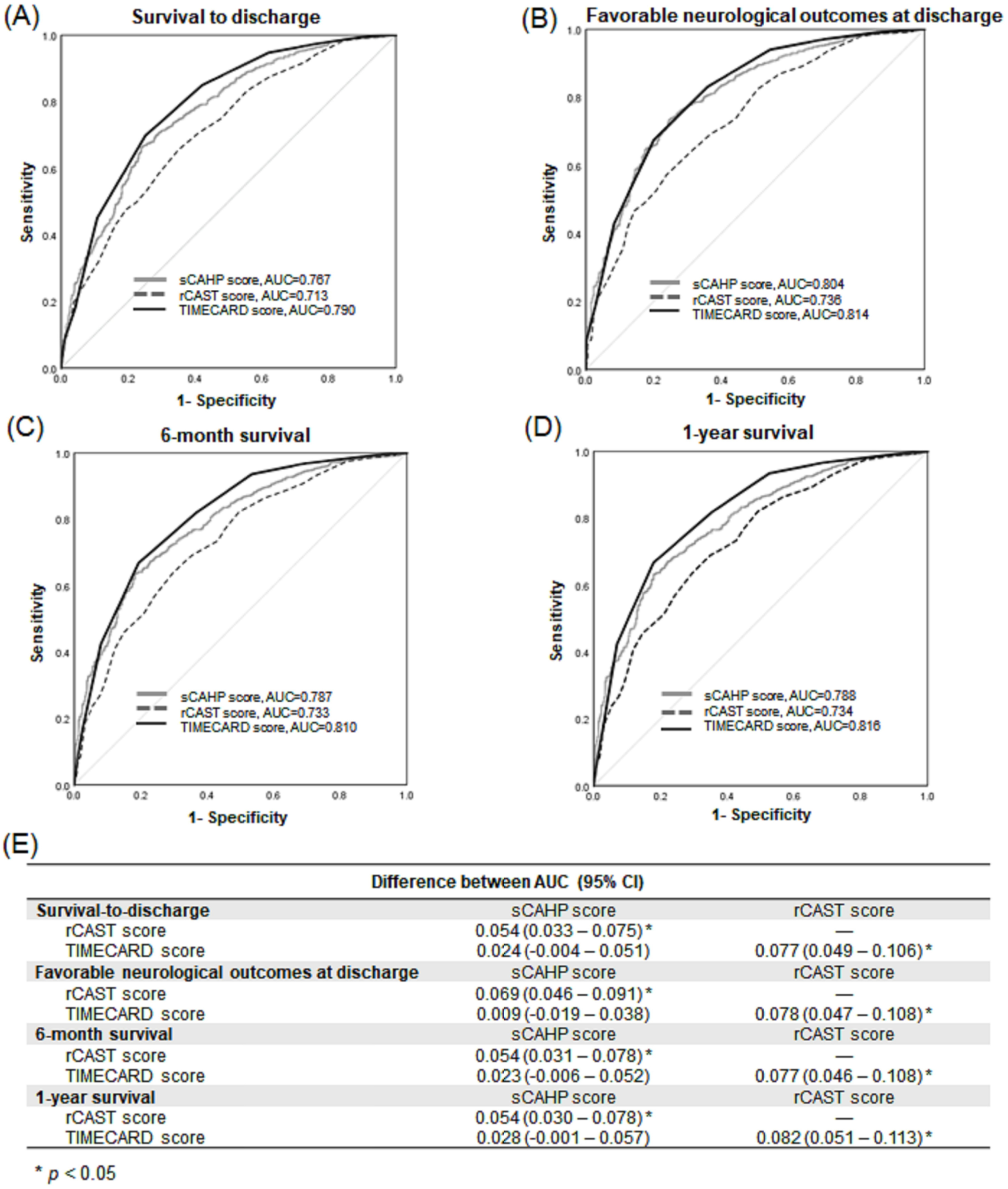
The ROC curves for enrolled patients. (A) Survival to discharge. (B) Favorable neurological outcomes at discharge. (C) 6-month survival. (D) 1-year survival. (E) Pairwise comparisons of AUCs for different severity scores. AUC: area under the ROC curve; CI: confidence interval; rCAST score: revised post-Cardiac Arrest Syndrome for Therapeutic hypothermia (CAST) score; sCAHP score: simplified Cardiac Arrest Hospital Prognosis (CAHP) score; TIMECARD score: TaIwan network of targeted temperature ManagEment for CARDiac arrest (TIMECARD) score; ROC curve: receiver operating characteristic curve.

The predictive ability of the severity scores was evaluated in subgroups stratified by age (65 years), initial shockable rhythms, site of CA (OHCA or IHCA), and pre-arrest neurological function. The ROC curves for long-term outcomes and pairwise AUC comparisons for each subgroup are presented in Supplementary Figure 2 (age), Supplementary Figure 3 (initial shockable rhythms), Supplementary Figure 4 (site of CA), and Supplementary Figure 5 (pre-arrest neurological function). The TIMECARD score demonstrated better discriminatory performance for 1-year survival in younger patients, those with IHCA, and those with good pre-arrest neurological status.

## Discussion

This retrospective single-center observational study enrolled 1773 adult patients with non-traumatic CA who were successfully resuscitated and demonstrated that the sCAHP, rCAST, and TIMECARD scores had a moderate-to-good ability to predict long-term outcomes in CA survivors. The sCAHP and TIMECARD scores demonstrated superior predictive ability for long-term outcomes compared with the rCAST score. Furthermore, the subgroup analyses indicated that the TIMECARD score had higher predictive ability for long-term outcomes than the sCAHP and rCAST scores in younger patients, those with IHCA, and those with good pre-arrest neurological status.

The superior ability of the sCAHP (4, 7–9) and TIMECARD (6) scores in predicting long-term outcomes compared with that of the rCAST score (5, 9, 10) could be attributed to various reasons. All three scoring systems incorporate similar variables such as initial rhythm, low-flow time, and pH levels. The sCAHP and TIMECARD scores also consider the site of CA (in public or at home) and bystander CPR, both of which are potentially associated with timely and prompt resuscitation for CA patients. Furthermore, the TIMECARD score includes call-to-start CPR duration, a metric similar to no-flow time. These resuscitation-related variables have been reported to be strongly associated with the prognosis and long-term outcomes of CA survivors (13–15). Variations in patient populations, prehospital emergency medical services, and post-arrest care interventions, including TTM and intensive care treatments, among the original studies that developed these scores may also affect their accuracy in predicting long-term outcomes for CA survivors.

Pre-arrest comorbidities and post-arrest functional status at discharge might be associated with the long-term outcomes of CA survivors. Amacher SA et al. demonstrated that favorable neurological status at discharge was linked to long-term survival (15). Chan PS et al. indicated that OHCA patients with more comorbidities were less likely to survive to discharge, and also reported that post-discharge mortality among OHCA survivors initially increased abruptly (27.0% at 3 months) and then rose gradually over time (37.1% at 1 year, 44.5% at 2 years, and 50.1% at 3 years) (25). These findings suggest that CA patients who survive beyond hospital discharge are more likely to experience a favorable long-term prognosis. Moreover, the superior predictive ability of the TIMECARD score over the sCAHP score was not initially observed at hospital discharge but became obvious as the follow-up continued, along with the proportion of patients aged below 65 years and IHCA increased, thereby revealing the predictive advantage of the TIMECARD score. This similar trend of the superior predictive ability of the TIMECARD score was also observed in patients with good prearrest neurological status.

The sCAHP, rCAST, and TIMECARD scores were selected to evaluate their predictive ability for long-term outcomes in CA survivors because of their rapid and straightforward variable acquisition after ROSC. However, because the sCAHP and rCAST scores omit some variables from their original formulas, their prognostic performance might be inferior to that of the TIMECARD score. Moreover, the TIMECARD score was developed using Taiwanese databases. Given that the current study was performed in a hospital in Taiwan, the TIMECARD score may have exhibited higher predictive accuracy than the other two severity scores.

An increasing number of hospitals and regions have recently published prediction models and severity scores for prognostication, utilizing artificial intelligence (AI) and machine learning methods. Several severity scores have been developed using machine learning, driven by recent advancements in AI (26–29). For research on long-term prognosis in CA survivors, the continued development of AI and machine learning methods is expected to improve the accuracy of prediction models and severity scores, enhancing their clinical applicability in the future.

This study has several limitations. First, its retrospective observational design makes selection bias unavoidable. In addition, the variables, prediction models, or severity scores not listed in the study could not be assessed for their ability to prognosticate long-term outcomes in CA survivors. Furthermore, because of variations in patients’ characteristics, accessibility and implementation of resuscitation, and differences in post-arrest care and management across various health-care systems, institutions, and countries, further prospective studies are needed to externally validate the generalizability of our findings. Besides, these severity scores were originally developed for early post-arrest assessment. This study sought to assess their utility in predicting long-term prognosis; however, the findings indicated that their predictive performance was less robust. Future investigations might consider developing a scoring system specifically tailored for the prediction of long-term outcomes following hospital discharge.

## Conclusion

The sCAHP, rCAST, and TIMECARD scores demonstrated good discrimination and calibration in predicting long-term outcomes in CA survivors. The TIMECARD score exhibited superior discriminatory performance for 1-year survival compared to the other two scores, particularly in younger patients, those with IHCA, and those with good pre-arrest neurological function.

## Data Availability

The datasets used and/or analyzed during the current study are available from the corresponding author on reasonable request.

## Declarations

### Ethics approval and consent to participate

The study was approved by the Institutional Review Board (IRB) of National Taiwan University Hospital (IRB number: 202407049RIND), and the requirement for informed consent was waived.

### Consent for publication

Not applicable.

### Authors’ contributions

All authors contributed to the study concept and design; Lin JJ, Chen WT, Lee HY, Ong HN, and Tsai MS contributed to the acquisition of the data; Tsai MS and Lin JJ analyzed and interpreted the data; Tsai MS and Lin JJ draft the manuscript; Huang CH, Chen WJ, and Chang WT provided critical revision of the manuscript for important intellectual content; Tsai MS and Lin JJ performed the statistical analysis; Huang CH and Chen WJ supervised the study. All authors read and approved the final manuscript.

## Acknowledgements

None.

## List of abbreviations

AUC: area under the receiver operating characteristic curve
CA: cardiac arrest
CAHP score: Cardiac Arrest Hospital Prognosis score
CAST score: post-Cardiac Arrest Syndrome for induced Therapeutic hypothermia score
CI: confidence interval
CPC score: cerebral performance category score
CPR: cardiopulmonary resuscitation
ED: emergency department
IHCA: in-hospital cardiac arrest
IRB: Institutional Review Board
NTUH: National Taiwan University Hospital
OHCA: out-of-hospital cardiac arres
rCAST score: revised CAST score
ROC curve: receiver operating characteristic curve
ROSC: return of spontaneous circulation
sCAHP score: simplified CAHP score
TIMECARD score: TaIwan network of targeted temperature ManagEment for CARDiac arrest score
TTM: targeted temperature management

---

Supplementary Figure 1. The Hosmer-Lemeshow goodness-of-fit test for enrolled patients.

(A) sCAHP score (6-month survival). (B) sCAHP score (1-year survival). (C) rCAST score (6-month survival). (D) rCAST score (1-year survival). (E) TIMECARD score (6-month survival). (F) TIMECARD score (1-year survival). rCAST score: revised post-Cardiac Arrest Syndrome for Therapeutic hypothermia (CAST) score; sCAHP score: simplified Cardiac Arrest Hospital Prognosis (CAHP) score; TIMECARD score: TaIwan network of targeted temperature ManagEment for CARDiac arrest (TIMECARD) score.

Supplementary Figure 2. The ROC curves for long-term outcomes stratified by age. (A) 6-month survival for patients aged ≤ 65 years. (B) 6-month survival of patients aged > 65 years. (C) 1-year survival for patients aged ≤ 65 years. (D) 1-year survival for patients aged > 65 years. (E) Pairwise comparisons of AUCs for different severity scores.

AUC: area under the ROC curve; CI: confidence interval; rCAST score: revised post-Cardiac Arrest Syndrome for Therapeutic hypothermia (CAST) score; sCAHP score: simplified Cardiac Arrest Hospital Prognosis (CAHP) score; TIMECARD score: TaIwan network of targeted temperature ManagEment for CARDiac arrest (TIMECARD) score; ROC curve: receiver operating characteristic curve.

Supplementary Figure 3. The ROC curves for long-term outcomes stratified by initial shockable rhythms. (A) 6-month survival for patients with initial non-shockable rhythms. (B) 6-month survival for patients with initial shockable rhythms. (C) 1-year survival for patients with initial non-shockable rhythms. (D) 1-year survival for patients with initial shockable rhythms. (E) Pairwise comparisons of AUCs for different severity scores.

AUC: area under the ROC curve; CI: confidence interval; rCAST score: revised post-Cardiac Arrest Syndrome for Therapeutic hypothermia (CAST) score; sCAHP score: simplified Cardiac Arrest Hospital Prognosis (CAHP) score; TIMECARD score: TaIwan network of targeted temperature ManagEment for CARDiac arrest (TIMECARD) score; ROC curve: receiver operating characteristic curve.

Supplementary Figure 4. The ROC curves for long-term outcomes stratified by the site of cardiac arrest (OHCA/IHCA). (A) 6-month survival for patients with OHCA. (B) 6-month survival for patients with IHCA. (C) 1-year survival for patients with OHCA. (D) 1-year survival for patients with IHCA. (E) Pairwise comparisons of AUCs for different severity scores.

AUC: area under the ROC curve; CI: confidence interval; IHCA: in-hospital cardiac arrest; OHCA: out-of-hospital cardiac arrest; rCAST score: revised post-Cardiac Arrest Syndrome for Therapeutic hypothermia (CAST) score; sCAHP score: simplified Cardiac Arrest Hospital Prognosis (CAHP) score; TIMECARD score: TaIwan network of targeted temperature ManagEment for CARDiac arrest (TIMECARD) score; ROC curve: receiver operating characteristic curve.

Supplementary Figure 5. The ROC curves for long-term outcomes stratified by pre-arrest CPC status. (A) 6-month survival for patients with poor pre-arrest CPC status. (B) 6-month survival for patients with good pre-arrest CPC status. (C) 1-year survival for patients with poor pre-arrest CPC status. (D) 1-year survival for patients with good pre-arrest CPC status. (E) Pairwise comparisons of AUCs for different severity scores.

AUC: area under the ROC curve; CI: confidence interval; CPC: cerebral performance category; rCAST score: revised post-Cardiac Arrest Syndrome for Therapeutic hypothermia (CAST) score; sCAHP score: simplified Cardiac Arrest Hospital Prognosis (CAHP) score; TIMECARD score: TaIwan network of targeted temperature ManagEment for CARDiac arrest (TIMECARD) score; ROC curve: receiver operating characteristic curve.

**Supplementary Table 1.** The variables of severity scores.

| Scores | sCAHP score | rCAST score | TIMECARD score |
| --- | --- | --- | --- |
| Variables | Age | Initial rhythm | Malignancy |
|  | Arrest settings (Home) | Witness to ROSC time | No bystander CPR |
|  | Initial rhythm | pH value | Non-shockable rhythm |
|  | Low-flow time | Serum lactate | Call-to-start CPR duration > 5min |
|  | Arterial pH value | GCS motor response score | CPR duration > 20min |
|  | Adrenaline dosage during CPR |  | Sodium bicarbonate use during CPR |
|  |  |  | GCS M1 at ROSC |
|  |  |  | No emergent CAG |

CAG: coronary angiography; CPR: cardiopulmonary resuscitation; GCS: Glasgow coma scale; rCAST score:
revised post-Cardiac Arrest Syndrome for Therapeutic hypothermia (CAST) score; sCAHP score:
simplified Cardiac Arrest Hospital Prognosis (CAHP) score; ROSC: return of spontaneous circulation;
TIMECARD score: Taiwan network of targeted temperature Management for CARDiac arrest (TIMECARD)
score.

